# How executive control and emotional reactivity influence coping strategies in psychiatric patients during the COVID-19 pandemic

**DOI:** 10.1101/2024.01.08.24300980

**Authors:** Josina D. Kist, Linda Schlüter, Fleur Duyser, Peter C.R. Mulders, Janna N. Vrijsen, Rose M. Collard, Philip F.P. van Eijndhoven, Indira Tendolkar

## Abstract

**Background:** During times of environmental challenges, adaptive coping strategies are essential to maintain mental health. Coping relies on executive control, which is often impaired in individuals with psychiatric disorders. Furthermore, emotional reactivity may interfere with executive control. Studying the association between cognitive skills and adaptive coping strategies, as well as the potential impact of emotional reactivity, could inform how we can provide mental support during large-scale adversity. In this study we examined coping strategies in a thoroughly phenotyped psychiatric cohort, the MIND-Set cohort, during the early COVID-19 pandemic stage.

**Methods:** We studied 1) the association between coping and both subjective and objective executive control before the pandemic, and three different coping strategies used during the pandemic, 2) the mediating role of emotional reactivity, indexed by amygdala reactivity, and 3) the moderating role of the presence of a psychiatric diagnosis in these associations. After finding no specific impact of patient or control status in this association, we decided to post-hoc study the transdiagnostic impact of depression severity in these associations.

**Results:** showed 1) only a significant association between subjective executive control and a self-reported positive reappraisal style and corona-related reappraisal. However, after controlling for depression severity, this association was no longer significant. Additionally, objective executive control was only directly associated with right amygdala reactivity, while amygdala reactivity in neither of the hemispheres mediated the association between executive control and any of the coping styles. Furthermore, the type of diagnosis did not moderate the association between executive control and coping.

**Conclusion:** Our findings firstly underline the difference between self-reported and performance based executive control. While both deficits in subjective and performance based EC may play a role in the persistence of psychiatric symptomatology, this finding emphasizes how depressive symptoms or negative affect can impact reappraisal ability. As this ability is fundamental to staying resilient, treatments focused on reducing negative affect and thereby training reappraisal are pivotal in the maintenance of mental health in the entire population during environmental challenges.

**Competing Interest Statement:** The authors have declared no competing interest.

## Introduction

Adopting adaptive coping strategies is essential for maintaining mental health during stressful situations, especially during broad environmental challenges such as the COVID-19 pandemic [1-4]. The maintenance or quick recovery of mental health after and during adversity indicates an individuals’ resilience, which needs active adaptation to a stressor through coping [5, 6]. Coping strategies are formed through an interplay of cognitive, emotional and behavioral processes in the attempt to manage adversity. These include strategies such as appraisal-, behavioral- and emotion-focused coping [7] Unsurprisingly, the use of these strategies is often altered in patients with psychiatric disorders [8-10]. Maladaptive coping may enhance symptom severity and overall stress susceptibility in patients with severe psychiatric comorbidity [11]. Likewise, pre-existing psychiatric morbidity prior to an environmental challenge may affect coping strategies and it may therefore be beneficial to estimate the relationship beforehand. This will be of specific relevance during specific environmental challenges such as the COVID-19 pandemic.

Coping strategies rely on executive control (EC) [12, 13], which describes cognitive processes that are required for the selection and monitoring of goal-directed behaviors, such as planning and organizing, and is instrumental in emotion regulation and the employment of adaptive coping strategies [14]. In healthy subjects, higher EC has been associated with the use of an adaptive type of coping called cognitive reappraisal [15]. Lower levels of EC, including deficits in inhibition, flexibility, working memory, planning, and global EC, has been associated with avoidance/disengagement and emotion-focused coping in different types of patient samples [16-18]. Patients with different psychiatric morbidity show global EC and working memory deficits [19, 20]. EC can be measured from an objective and subjective point of view, whereby the correlations between both types of measurements have been found to be weak to moderate [21, 22]. Objective EC can be quantified through performance-based tasks that are regarded as a more direct measure of EC. However their ecological validity has been questioned [23]. Subjective, or self-reported, measures of EC on the other hand are highly susceptible to negative affect and individual personality traits and may thus show a different relationship with coping strategies [21]. Self-reported EC deficits have been shown to exhibit a stronger correlation with depressive symptoms, potentially through the influence of negative self-referential processes in evaluations of cognitive performance [24, 25]. While self-report EC may indicate presence of a so-called Cognitive Impairment bias, this measure should not be disregarded [24]. Both deficits in subjective and performance based EC may play a role in the persistence of psychiatric symptomatology, as these indicate negative information processing biases and may impair emotional control [26].

Emotion-focused coping is dependent on emotional reactivity to negative information, which may be elevated when top-down emotional control is lacking [27]. A neural indicator for emotional reactivity is the level of amygdala activity, which signifies sensitivity of the amygdala in response to emotional information. A stronger amygdala reactivity is linked to diminished top-down EC and is commonly observed in a wide range of psychiatric disorders, including mood, anxiety, and neurodevelopmental disorders [28-30]. Consequently, heightened amygdala reactivity to emotionally salient information may override top down EC and promote a reliance on less adaptive coping strategies.

Despite a large body of evidence highlighting the mental health consequences of the COVID-19 pandemic, including how pre-existing EC deficits may impact health-related behavior, it remains unclear how different psychiatric conditions and diminished EC impact thoughts and coping behaviors related to the pandemic [31-33]. Since coping strategies are found to be related to both EC and emotional reactivity, the covid-pandemic presents an opportunity to study specific stress-related coping styles in a large naturalistic psychiatric patient cohort in a transdiagnostic manner. In this study we therefore assess coping behavior in a well-phenotyped and naturalistic outpatient sample with neurodevelopmental (ADHD, ASD) and/or stress-related disorders (i.e. depression, anxiety and addiction) and a participant sample without psychiatric disorders during the early months of the covid-pandemic.

We conceptualized coping in three different ways to account for various relevant strategies and following the approach previously adopted by other studies using the DynaCore during the covid pandemic [33]. The three coping techniques measured were positive appraisal style (PAS), corona-related reappraisal (CRR) and behavioral coping style (BCS). PAS and CRR represent forms of cognitive reappraisal which describes a re-evaluation to adapt a first impression of a situation, thereby altering the emotional and physiological experience, without having an unrealistic positive perception [34]. Behavioral coping describes the way one actively uses adaptive strategies to handle a stressful situation, such as seeking support methods, planning steps to counteract adversity, and the use of venting and humor. Both these types of coping have been found useful, since higher indications of using these strategies is related to lower negative affect in general, but also specifically for patients to cope with their mental illness by reframing experiences in a more positive light and actively adjusting their behavior/social position [35, 36].

We aimed to investigate 1) whether the three different coping strategies during the COVID-19 pandemic were related to different forms of EC (subjective/objective) and 2) the mediating role of emotional reactivity to negative stimuli prior to the start of the pandemic. Furthermore we assessed 3) whether the presence of any psychiatric diagnosis before the COVID-19 pandemic influenced the association between EC and coping. Therefore, this study takes a transdiagnostic approach in assessing coping and EC impairments, and may thus further inform measures on how to keep citizens with and without psychiatric disorders resilient during environmental challenges.

## Results

The selection of the MIND-Set cohort included in this study consisted of 88 patients and 49 controls. Demographics and depressive symptom severity were measured both before (T0) and during the covid pandemic (T1). The objective and subjective measures of EC and emotional reactivity were collected only during T0, while the coping strategies were only measured at T1 (Table 1, see Figure 1 for the timeline of data collection).

**Table 1.** Descriptive information of demographics, depressive symptom severity, measures of executive control, emotional reactivity and coping strategies of the patient (n = 88) and control (n = 49) sample before (T0) and during the covid pandemic (T1). Abbreviations: IDS-SR: Inventory of Depressive Symptomatology-Self Report; GEC: Global Executive Control

|  | T0 |  | T1 |  |
| --- | --- | --- | --- | --- |
|  | Patients | Controls | Patients | Controls |
| Age in years, mean (SD) | 38.60 ($\pm 13.96$) | 37.90 ($\pm 17.10$) | 41.08 ($\pm 14.12$) | 40.67 ($\pm 16.12$) |
| Gender (%) |  |  |  |  |
| Male | 54.5 | 40.8 | 54.5 | 42.9 |
| Level of education (%) |  |  |  |  |
| None | 1.1 | 0.0 | 1.1 | 0.0 |
| Low | 19.1 | 6.1 | 10.2 | 2.0 |
| Medium | 37.5 | 34.7 | 27.3 | 18.4 |
| High | 42.0 | 59.2 | 61.4 | 79.6 |
| Depressive Symptom Severity<br>IDS-SR (SD) | 33.86 ($\pm 13.17$) | 4.90 ($\pm 3.83$) | 23.94 ($\pm 14.76$) | 7.59 (10.84) |
| Executive control |  |  |  |  |
| BRIEF-A |  |  |  |  |
| GEC (SD) | 71.93 ($\pm 4.05$) | 46($\pm 4.08$) | - | - |
| Spatial Working Memory Task |  |  |  |  |
| Strategy (SD) | 31.94 ($\pm 5.48$) | 31.80 ($\pm 6.10$) | - | - |
| Between Errors (SD) | 23.06 ($\pm 19.46$) | 18.71 ($\pm 17.26$) | - | - |
| Emotional Reactivity |  |  |  |  |
| Left Amygdala (SD) | 0.31 ($\pm 0.23$) | .34 ($\pm 0.25$) | - | - |
| Right Amygdala (SD) | 0.30 ($\pm 0.20$) | 0.28 ($\pm 0.24$) | - | - |
| Coping |  |  |  |  |
| Positive Appraisal Style (SD) | - | - | 0.40 ($\pm .15$) | 0.54 ($\pm .14$) |
| Corona Related Reappraisal SD) | | | 5.72 ($\pm 1.64$) | 6.69 ($\pm 1.49$) |
| Behavioral Coping (SD) | | | 19.64 ($\pm 4.04$) | 21.04 ($\pm 4.16$) |

**Table 2.** Association Between Executive Control and Coping.

|  | PAS |  |  | CRR |  |  | BCS |  |  |
| --- | --- | --- | --- | --- | --- | --- | --- | --- | --- |
| | *B* | $\beta$ | *p* | *B* | $\beta$ | *p* | *B* | $\beta$ | *p* |
| Subjective EC |  |  |  |  |  |  |  |  |  |
| GEC | -.003 | -.274 | .003* | -.028 | -.266 | .002* | -.030 | -.111 | .174 |
| Objective EC |  |  |  |  |  |  |  |  |  |
| Between Errors | -.001 | -.078 | .900 | -.004 | -.045 | .900 | -.021 | -.096 | .900 |
| Strategy | -.001 | -.020 | 1.000 | -.001 | -.005 | 1.000 | -.017 | -.024 | 1.000 |

*Note.* Linear regression analysis testing the association between coping (Positive Appraisal Style [PAS], Behavioral Coping Scale [BCS], Corona-Related Reappraisal [CRR]). Higher scores indicate more frequent coping or reappraisal use. Means were adjusted for gender, age, and the level of education. Range of the coping outcome variables: PAS 0-1, BCS 8-32, CRR 2-10, Emotion- Focused Coping 6-24, Problem-Focused Coping 4-16

**Figure 1.**
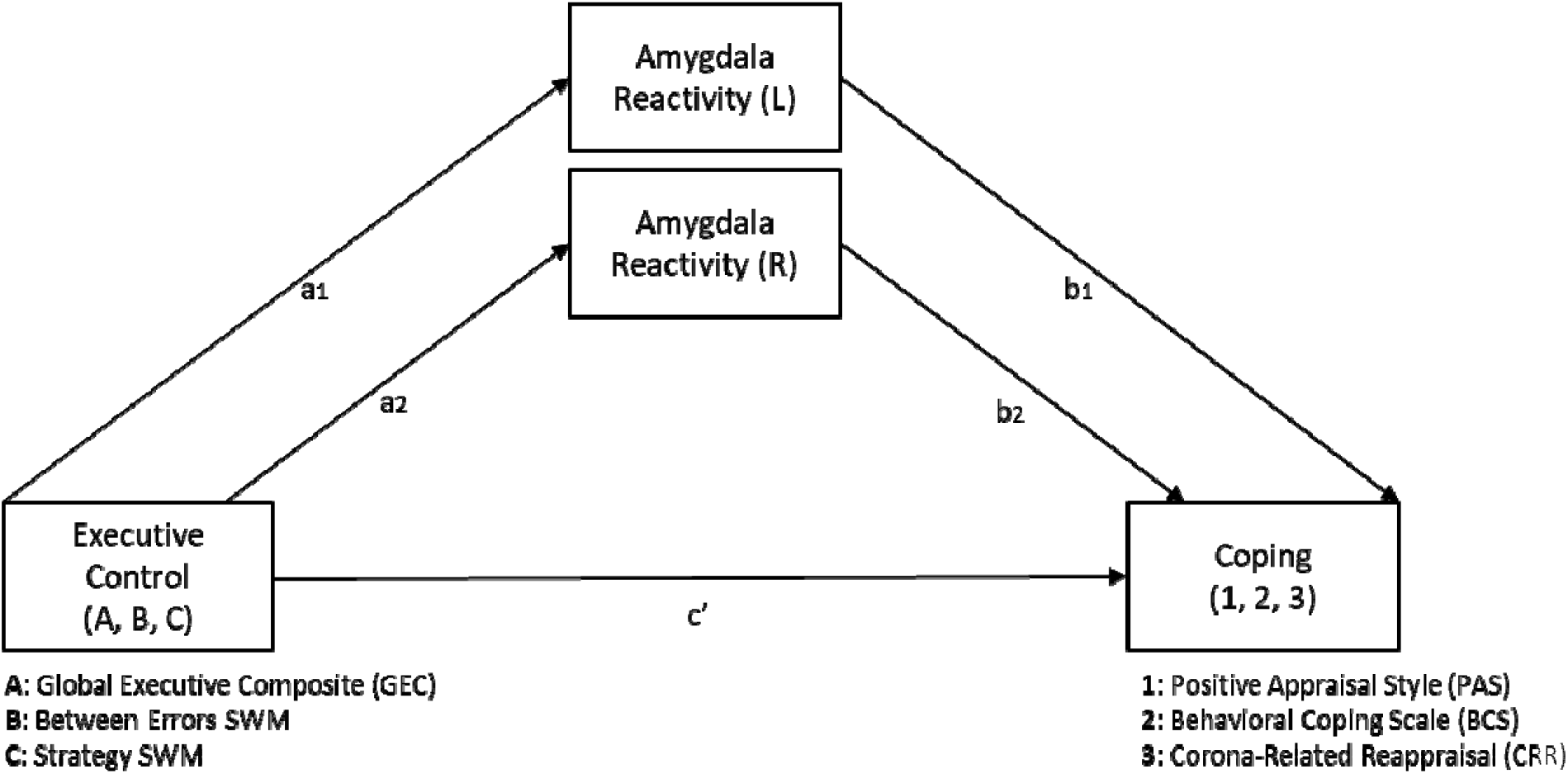
The Mediating Effect of Amygdala Reactivity on the Association Between Executive Control and Coping. *Note*. Mediation model illustrating the direct effect of executive control (Global Executive Composite score from the BRIEF-A, Between Errors and Strategy scores from the Spatial Working Memory [SWM] task) on coping (Positive Appraisal Style, Behavioral Coping Scale, Corona-Related Reappraisal) and the indirect effect via left and right amygdala reactivity. a1, a2, b1, b2 and c’ indicate path coefficients.

### General group differences between patients and controls at T0 and T1

Examination of demographic differences at baseline showed that the patient and the control group did not differ in age (*t*[135] = -.26, *p* = .795 at T0, *t*[135] = -.15, *p* = .878 at T1), gender (χ2[1] = 2.37, *p* = .123 at T0, χ2[1] = 1.47, *p* = .226 at T1), and the level of education (χ2[3] = 6.30, *p* = .098 at T0, χ2[3] = 6.02, *p* = .110). The patient group reported a significantly higher level of depressive symptomatology than the control group both at T0 and at T1 (t[109.3] = -19.13, *p* < .001). The IDS score for patients at T0 was significantly higher than at T1 (t[df=86]=6.45, p <.001), while for controls the difference for IDS score was not significant between T0 and T1 (t[df=48]= -1.91, p = 0.06)

Comparison of the objective EC measures showed that there were no significant group differences for the SWM Strategy and the Between Error scores (*F*[2,128] = 1.21, *p* = .30, ηp^2^ = .019). Regarding the subjective EC measures, the BRIEF-A at T0, the multivariate test revealed that patients scored significantly higher on all measures than the control group (*F*[12,118] = 22.30, *p* < .001), with a large effect size (ηp^2^ = .69). There was no significant difference in left or right amygdala reactivity between the patient and the control group neither before nor after controlling for stress ratings prior to the task (*F*[2,127] = .13, *p* = .559, ηp^2^ = .009).

While there were no group differences in the behavioral coping scale (BCS), the patient group reported significantly lower positive appraisal style (PAS) and corona-related reappraisal CRR scores than the control group (F[1,129]=20.65, p< 0.001, with a large effect size of ηp^2^ = .14 and F[1,129]=8.95, p <0.05, with a medium effects size of ηp^2^ = 0.065, respectively), indicating that at the beginning of the COVID-19 pandemic patients made less frequent use of positive and corona related reappraisal.

### The Association Between Executive Control before and Coping strategies during the pandemic

Coping was differently associated with self-reported EC as compared to performance-based EC. The GEC was significantly negatively associated with PAS (F[1,136] = 4.05, p = <0.001, R^2^ = 0.18) and CRR (F[1,136] = 2.99, p = <0.001, R^2^ = 0.14), but not with BCS (F[1,136] = 4.97, p = <0.001, R^2^ = 0.21). Thus, higher levels of self-reported executive dysfunction were related to less frequent PAS and CRR.

## The Mediating Effect of Amygdala Reactivity

When testing the mediating effect of amygdala reactivity on the association between the three different EC variables (GEC/Between Errors and Strategy) and the different coping styles (PAS/BCS/CRR) we only found a significant association between the Between Error scores from the SWM task and right amygdala reactivity (*B* = .003, β = .254, *p* = .03), suggesting worse SWM performance was associated with an increase in right amygdala reactivity (Table S2 in the Supplementary materials).

## The Impact of Psychiatric Diagnosis and Depressive Symptom Severity on the Relation Between Executive Control And Coping

### The impact of a Psychiatric Diagnosis

A moderation analysis was conducted to test whether a psychiatric diagnosis impacted the association between EC and coping strategies. A significant moderation effect of group was observed only for the association between the self-reported GEC score and BCS (*F*[1,127] = 9.25, *p* = .009, *R*2 change = .05; Table S3 in Supplementary materials). In the control group, higher levels of GEC scores, indicating lower EC, was significantly associated with an increase in BCS (*B* = .213, *p* = .016). In the patient group, the opposite effect was observed: an increase in GEC was significantly associated with a decrease in BCS (*B* = -.074, *p* = .048).

### Post-Hoc: The Impact of Depressive Symptom Severity

Only self-reported EC was found to be associated with two types of coping strategies. Since earlier literature indicated that variance in self-reported or subjective EC may be caused by symptoms of depression, we subsequently conducted analyses to determine whether the severity of depressive symptoms was an explanatory variable in this relationship [24, 25]. The IDS-SR score was used to measure depressive symptom severity both at T0 and T1. Symptom severity at T0 was used as a covariate to assess the impact of depressive symptom severity firstly on the group differences between patients and controls in EC measures, secondly on the association between EC and coping strategies and thirdly on the moderation of patients vs controls in the association between EC and coping.

The significant group differences on subjective EC scores between patients and controls remained significant when controlling for depressive symptomatology using the IDS-SR total score at T0, (supplementary table S1).

After adding the depression severity scores as a covariate in the linear regressions, the previously found significant associations between the subjective EC (GEC) and the PAS and CRR were not significant anymore (Table 3.)

**Table 3.** Impact of controlling for Depressive Symptom Severity (IDS-SR) at T0 on Association Between Executive Control and Coping, and Coping Measures.

|  | PAS |  |  | CRR |  |  | BCS |  |  |
| --- | --- | --- | --- | --- | --- | --- | --- | --- | --- |
| | *B* | $\beta$ | *p* | *B* | $\beta$ | *p* | *B* | $\beta$ | *p* |
| GEC | -.003 | -.274 | .003* | -.028 | -.266 | .002* | -.030 | -.111 | .174 |
| **GEC + IDS-SR** | **.000** | **.037** | **.773** | **.008** | **.077** | **.552** | **-.010** | **-.036** | **.780** |

Note. Linear regression analysis on the association between executive control (Global Executive Composite [GEC] scores of the BRIEF-A and coping (Positive Appraisal Style [PAS], Corona-Related Reappraisal [CRR], and the Behavioral Coping Scale [BCS]). For the first model with Gender, age, and the level of education were entered as covariates ( $N = 137$ ), for the second model using the Depressive symptomatology (IDS-SR) at T0 as additional covariate ( $N=137$ ). Higher GEC scores indicate a greater level of dysfunction.
\* significant at the .05 level, \*\* significant at the .001 level

The association between GEC scores and BCS in the patient group was also affected by the severity of depressive symptomatology. Additional post-hoc analyses were carried out that can be found in part 3 of the supplementary materials and reveal that the group differences were no longer significant.

Altogether, we no longer observe a significant association between EC and coping strategies, and between group, EC and coping strategies, when controlling for depressive severity.

## Discussion

In this study we related coping strategies during the COVID-19 pandemic to different forms of EC (subjective/objective) and/or emotional reactivity in comorbid psychiatric patients prior to the pandemic. Though objective EC measures did not discriminate between patients and controls, a negative relationship between executive function and amygdala activity as a proxy of emotional reactivity was confirmed. Comparison of subjective EC measures at baseline revealed significant differences between patients and controls even when controlling for depressive symptom severity. Higher levels of self-reported executive dysfunction before the covid pandemic was related to less frequent positive appraisal style (PAS) and covid-related reappraisal (CRR) during the pandemic. However, when controlling for depressive symptoms at baseline, these effects no longer were significant. Below we will discuss the potential implications of these findings

### Group Differences

Surprisingly, we did not find any differences in objective EC measures between patients and controls. Several factors could account for this observation. Firstly, EC may be relatively well preserved in this outpatient cohort which does not exhibit specific neurocognitive problems, leading to a potential ceiling effect in EC task performance. While previous studies have demonstrated a good validity of the SWM task, indicating both case-control differences as well as associations with a negative attributional style, the ecological validity of neurocognitive tests may also be called into question [37-39].

Secondly, the lack of differences between patients and healthy controls may be explained by the large heterogeneity of this patient cohort. However, this outpatient cohort judged their executive performance significantly worse than the control group even when controlling for depressive symptoms. This significant difference in subjective EC was found to be independent of depressive symptom severity.

Earlier research has indicated that there is a large discrepancy between outcomes of subjective and objective measures of EC, which may be due to methodological and conceptual differences [40]. While self-report measures of EC are often critiqued due to their vulnerability to depressive symptomatology, these reports do indicate everyday performance and underline the importance of the affective state therein. Overall, in assessing EC, the method through which this is done needs to be carefully considered and reported.

### Association EC and Coping

In the relationship between EC and coping, we identified a significant association between subjective EC and positive appraisal style (PAS), as well as subjective EC and corona related reappraisal (CRR). However, upon including depression severity in the analysis, these associations were no longer significant. This highlights the impact of the affective state on coping strategies, particularly the use of cognitive reappraisal. Furthermore, the stronger association between coping strategies and subjective EC rather than performance-based EC may be attributed to the use of self-report questionnaires for these concepts. Later in this discussion, we will touch upon the impact of the affective state on coping and self-report more in-depth.

The absence of a significant association between performance on the SWM task and any of the coping strategies might be explained by two potential reasons. Firstly, the hypothesized relationship between EC and coping strategies might not exist in this specific sample. Alternatively, the SWM task may not capture EC validly in this study. As explained above, the variability in SWM task performance might not accurately represent the variability in EC usually seen in a patient population [37].

### Mediating role of Emotional Reactivity

In the absence of objective EC changes, it is not surprising that we observed no differences in amygdala reactivity between patient and controls, since this measure of emotional reactivity is proposed to be affected by top down cognitive control. We did, however, identify a general pattern where poorer SWM performance was associated with an increase in right amygdala reactivity. This finding supports the notion that a decrease in top-down control, as indicated by lower SWM performance, might facilitate a stronger emotional/affective bottom-up reaction [41-43]. Our findings corroborate previous reports pointing towards a link between working memory performance and amygdala reactivity, which, however, emphasized the left amygdala as a more sensitive predictor of WM performance [43]. Despite the observed association between SWM performance and amygdala reactivity, we conclude that the proposed mediation model cannot sufficiently explain differences in coping.

## Importance of the affective state

Self-reported coping abilities were most strongly influenced by depressive symptomatology, which also impacted the relationship between subjective EC at baseline and coping at follow up. Interestingly controlling for depressive symptomatology seemed to primarily impact PAS and CRR while the results regarding behavioral coping style (BCS) remain unaffected. This indicates that the effect of these symptoms is not tied to self-report measures of coping per se, but more so the ability to reappraise. Reappraisal, a type of coping and emotion regulation that entails the ability to look at a situation from a different perspective, requires both cognitive skill as well as positive affect [15, 34]. As reappraisal skills are vulnerable to impaired cognition and cognitive biases, while they are an important tool in remaining resilient. As such, this study highlights the impact of a negative affective state on self-report of EC and, importantly, on reappraisal style. However, it is noteworthy that the difference between the subjective EC scores between patients and controls persisted, even after controlling for age, gender, the level of education and depressive symptoms. Therefore, this study shows that while self-reported EC may be affected by the severity of depressive symptoms, there remains a difference between patients and controls in how they self-assess their EC. Stronger cognitive skills may play a role in mitigating the perpetuation of negative affect.

## Strengths and limitations

This study is one of the first to investigate the relationship between subjective and objective types of EC, coping and emotional reactivity in patients with different psychiatric conditions and as such can be evaluated to reflect transdiagnostic outcomes. The naturalistic and multimorbid nature of the patient cohort represents a strength of this study, as it facilitates the detection of transdiagnostic mechanisms underlying psychopathology (Krueger & Eaton, 2015; Insel, 2014). This further suggests that the ecological validity of subjective forms of EC can be seen independent of diagnosis.

However, this study also has limitations. Firstly, while the naturalistic cohort is one of the main strengths, it also poses the challenge of heterogeneity within this study. Secondly, we acknowledge the discrepancy of measurements during baseline and follow up: We lack coping measures before covid (T0), and did not assess emotion regulation during follow up (T1), preventing us from exploring longitudinal changes in coping styles, and the impact of emotion regulation on coping styles. Furthermore, the variable time that has passed between T0 and T1 between participants may influence the association between measurements (as participants were included for T0 over the span of four years from 2016 until 2020, while all were approached concerning their participation in the follow-up in august 2020.)

It is likely that environmental challenges, such as the COVID-19 pandemic or natural disasters related to climate change, will become more frequent. To protect and sustain mental resilience, a more in-depth understanding of how EC impacts coping strategies in both non-disordered individuals and those with one or several psychiatric diagnoses will be critical in promoting more adaptive coping strategies. Therapies that bolster executive control, potentially mitigating cognitive impairment bias, may prove advantageous in fostering the adoption of positive reappraisal techniques and other adaptive coping strategies, thereby contributing to overall psychological well-being.

## Methods

This study used data from the Measuring Integrated Novel Dimensions (MIND-Set) psychiatric cohort study that was initiated by the Department of Psychiatry of the Radboud university medical center (Radboudumc) and the Donders Institute in Nijmegen, the Netherlands [44]. Initial data collection (baseline, T0) started in 2016 as a cross-sectional study and included questionnaires assessing demographics and symptomatology, behavioral tasks, neuropsychological tests, and the collection of biometric material and neuroimaging measures [44]. With the surge of the COVID-19 pandemic and the by the government implemented restrictions in 2020, participants were approached for an online follow-up study which included additional questionnaires tailored to assess the effects of and coping behaviors in response to the pandemic between August 2020 and July 2021. In this study, we focused on data from baseline (T0) and a follow-up assessment (T1) that reflected the peak of the restrictions and threat due to absence of vaccination in the Netherlands.

## Participants

The participant sample consisted of 88 psychiatric patients (45.5% female, mean age 38.6 (±13.96) and 49 non-disordered matched controls (59.2% female, mean age 37.9 (±17.10)) (see Table 1). Patients were eligible for inclusion if they were clinically diagnosed with at least one stress-related (mood and/or anxiety disorders) or neurodevelopmental disorder (Attention-Deficit/Hyperactivity Disorder [ADHD] and/or autism spectrum disorder [ASD]) (see van Eijndhoven et al. 2022 for more detailed information on inclusion and exclusion criteria).

### Measures

At baseline (T0), before the COVID-19 pandemic, executive control (EC) and emotional reactivity were measured. At follow-up (T1), we assessed different coping strategies. Depressive symptom severity was measured both at T0 and T1 to account for the impact of negative affect on subjective measures (see Table 1).

## Executive Control Measures

### Spatial Working Memory Task

Objective EC was measured using the spatial working memory (SWM) task from the Cambridge Neuropsychological Test Automated Battery (CANTAB) [37, 45]. This test-battery has been used in many studies for assessment of a variety of psychiatric and non-disordered samples, and has shown to reliably detect variation in EC [37, 46-48]. We used two outcome measures indicating the maintenance and manipulation of visuospatial information: The *Between Errors* score indicates the number of mistakes made during the task and is more strongly related to working memory specifically. The *Strategy* score measures consistency in the approach of the task, reflecting more general executive functioning [49, 50]. Higher scores on both outcome measures indicate worse performance and decreased executive control.

### Behavior Rating Inventory of Subjective Executive Function–Adult Version (BRIEF-A)

Subjective EC performance was measured with the BRIEF-A. participants were asked to indicate how frequently they had experienced executive problems during their daily lives (e.g. “I have trouble making decisions”) within the past month on a 3-point Likert scale. From a total of 75 items, nine non-overlapping clinical scales are derived that form two broader indices: the Behavioral Regulation Index (BRI, comprising the scales Inhibit, Shift, Self-monitor and Emotional Control; range: 30 - 90), and a Metacognition Index (MI, comprising the scales Plan/Organize, Initiate, Task Monitor, Working Memory, and Organization of Materials; range: 40 - 120). The sum of both results in the Global Executive Composite (GEC; range: 70 - 210). Higher scores indicate increased executive dysfunction [51, 52].

### Amygdala Reactivity Measured by the Emotional Faces Processing Task at T0

Participants completed the emotional faces processing task during a functional MRI scan to assess the sensitivity of the amygdala in response to negative emotional information [53-56]. The task consists of two different conditions: the first in which the emotional expression of the upper face needs to be matched to either of two lower faces (the ‘face’ condition). Only negative emotional expressions (anger and fear) were included. In the ‘shape’ condition, faces were replaced with elliptical pixelated faces and matching was done with regards to spatial orientation. Participants completed two ‘face’ and three ‘shape’ blocks that consisted of six trials of 30s each. The fMRI data acquisition settings during the emotional faces processing task have previously been described in a different publication from the MIND-Set study [57]:

fMRI data were collected using a 3T Siemens Magnetom Prisma system with a 32-channel head coil. T2*-weighted echoplanar images with blood-oxygen-level-dependent contrast were acquired during the emotion processing task (repetition time [TR] = 1000 ms, echo time [TE] = 34 ms, slicing: interleaved ascending, voxel size: 2.0 × 2.0 × 2.0 mm, flip angle: 60°). Anatomical images were acquired using a T1-weighted MP-RAGE sequence (TR = 2300 ms, TE = 3.03 ms, voxel size: 1.0 × 1.0 × 1.0 mm, flip angle: 8°, GRAPPA acceleration factor: 2). [57]

Pre-processing and first- and second-level analysis of the fMRI data were carried out in SPM12 (Wellcome Department of Imaging Neuroscience, London, United Kingdom). A detailed description of the pre-processing parameters can be found in previous work by Duyser and colleagues [57]. Single-subject parameter estimates for the faces vs. shapes contrast, indicating regions of increased activity for the faces relative to the shapes, were obtained in a first-level analysis by means of a one-sample t-test. As an indicator of reactivity to negative emotional information, parameter estimates (beta weights) for the faces vs. shapes contrast were extracted from the left and right amygdala by using the MarsBaR toolbox with regions of interest from the Automatic Anatomical Labeling (AAL) atlas [57, 58].

### Inventory of Depressive Symptomatology – Self-Report (IDS-SR)

The IDS-SR was used to assess depressive symptomatology over the past two weeks [59]. It consists of 28 constant and two optional items that are rated on a 4-point Likert scale scored from 0-3, and a Cronbach’s alpha of 0.85 [59]. A higher rating indicates increased symptom severity.

## Measures at T1

### Coping

Coping was operationalized in three ways using self-report scales from the DynaCORE questionnaire. The DynaCORE was developed for the EU DynaMORE project (see www.dynamore-project.eu) and comprises a collection of pre-existing and newly developed scales related to resilience and experiences during the COVID-19 pandemic [60].

The following outcome measures of the DynaCORE were utilized within this project: Positive *Appraisal Style* (PAS), *Behavioral Coping Scale* (BCS), and *Corona-Related Reappraisal* (CRR). All three measures have previously been linked to increased resilience during the COVID-19 pandemic [60, 61]. Positive Appraisal Style (PAS) measures the general tendency towards a positive appraisal of stressors. A positive appraisal style is defined by the absence of negative biases while simultaneously refraining from an unrealistic or delusional positive appraisal [60]. Examples of items from this measure are “I tell myself that there are worse things in life” and “I think that the situation also has its positive sides”. As PAS is measured with items from different validated questionnaires with distinct rating scales [62, 63], raw scores were standardized using the ‘Proportion of Maximum Scaling’ (POMS) method to maintain the relative differences between the observed scores [64, 65]. The Behavioral Coping Scale (BCS) assesses typical thinking processes and behaviors in the face of challenges, with a focus on positive coping behavior [60]. It is measured with eight items rated on a 4-point Likert scale and raw scores are summed up to yield a total BCS score (range 8 - 32). Examples of items from this measure are “I try to come up with a strategy about what to do” and “I try to get advice or help from other people about what to do”.

Corona-Related Reappraisal (CRR) measures positive appraisal of the corona crisis specifically (eg. “I expect that I will learn something positive from of the Corona pandemic for my own life.”) and is assessed using the sum of two items on a 5-point Likert scale (range: 2 - 10).

For all measures, a higher score indicates a higher rate of the coping strategies.

## Statistical Analyses

All analyses were conducted in IBM SPSS Statistics 25. Mediation and moderation analyses were conducted using the PROCESS macro for SPSS [66]. Residuals were normally distributed and independent and homoscedasticity and multicollinearity diagnostics were within an acceptable range. Gender, age at T0, and the level of education at T0 were included as covariates when investigating the relationship between different forms of executive control and covid-related coping in later analyses.

### Group differences

Differences in the level of executive control (EC) and amygdala reactivity measured at T0 between patients and controls (CN) was tested using multivariate analyses of covariance (MANCOVA). This was also done for the three coping styles measured at T1: positive appraisal style (PAS), behavioral coping style (BCS) and covid-related reappraisal (CRR). Gender, age at T0, and the level of education at T0 were added as covariates. Differences were considered significant at p<0.05, and p-values were adjusted for multiple testing with a Holm-Bonferroni correction when required.

### The Association Between Executive Control And Coping

We used linear regression analyses to investigate the association of objective EC measures (Between Errors and Strategy scores of the SWM) and self-reported levels of EC, as measured by the Global Executive Composite (GEC) T-score of the BRIEF-A, with coping strategies. Separate analyses were carried out with each of the three coping measures (PAS, BCS and CRR) as outcome variables.

### The Mediating Role of Amygdala Reactivity

To assess whether the relation between EC and coping is mediated by amygdala reactivity, separate mediation analyses with the abovementioned self-report and performance-based EC measures (GEC T-scores, Between Errors, Strategy scores) as independent variables and coping measures as dependent variables (PAS, CRR, BCS) were performed (see Figure 1). The beta values obtained from the left and right amygdala were entered as mediating variables.

### The Impacting Presence of Psychopathology

To determine whether the relation between EC and coping was differently affected in psychiatric patients and healthy controls, the above mentioned linear regressions were repeated with group (patient/control) added as a moderator variable.

### Post-Hoc: Impact of Depressive Symptomatology

Additionally, as a Post-Hoc, we tested for the impact of depressive symptomatology at baseline by adding the IDS-SR at T0 1) to the MANCOVA when testing group differences 2) to the covariates of the linear regressions testing the associations between EC and coping 3) to the covariates of the linear regressions using patient/control status as a moderator variable.

## Data Availability

All data produced in the MIND-Set study are available upon reasonable request to the authors

