## Supplementary Materials for "How executive control and emotional reactivity influence coping strategies in psychiatric patients during the COVID-19 pandemic"

**Table S1.** Comparison of self-report based EC scores at baseline in the Patient and Control group also controlled for depression symptomatology (IDS-SR) at baseline.

|  | Adjusted Mean (SE) | | *F*(1,129) | ηp² |
| --- | --- | --- | --- | --- |
| **BRIEF-A** | Patients | Controls |  |  |
| Inhibition | 64.26 (4.98) | 53.97 (4.92) | 9.69* | .07 |
| Shift | 67.89 (4.51) | 56.06 (4.46) | 15.60** | .11 |
| Emotional Control | 57.70 (4.00) | 49.04 (3.95) | 10.66* | .08 |
| Self-Monitor | 57.38 (4.92) | 47.08 (4.86) | 9.94* | .07 |
| Initiate | 67.74 (4.90) | 48.59 (4.84) | 34.68** | .22 |
| Working Memory | 67.25 (3.93) | 55.28 (3.89) | 20.99** | .14 |
| Plan/Organize | 58.82 (4.78) | 45.18 (4.73) | 18.46** | .13 |
| Task-Monitor | 64.41 (4.71) | 53.33 (4.66) | 12.55* | .09 |
| Organization of Materials | 63.40 (5.29) | 50.29 (5.23) | 13.92** | **.**10 |
| Behavioral Regulation Index (BRI) | 63.36 (3.89) | 51.44 (3.84) | 21.31** | .14 |
| Metacognition Index (MI) | 66.94 (4.45) | 50.44 (4.40) | 31.11** | .20 |
| Global Executive Composite (GEC) | 66.36 (3.86) | 50.83 (3.81) | 36.78** | .23 |

Note. ANCOVAs comparing BRIEF-A scores between the patient group (N = 88) and the control group (N = 49). Higher scores on all BRIEF-A scales indicate greater dysfunction. Means were adjusted for gender, age the level of education and depressive symptomatology.
 * significant at the .05 level, ** significant at the .001 level

**Table S2**. The analysis on the mediating effect of amygdala reactivity on the association between executive control (EC) and coping Styles

| *Coping Style* | *EC Variable (IV)* | *Mediator* | *Effect of IV on mediator*  *(a1, a2)* | *Unique mediator effect (b1, b2)* | *Indirect effect (a, b)* | *Confidence Interval* |
| --- | --- | --- | --- | --- | --- | --- |
| **PAS** |  |  | *B*(β) | *B*(β) | *B*(β) | 95%CI |
|  | GEC | Left AR | .001 (.078) | .005 (.007) | .000 (.001) | [-.027, .027] |
|  |  | Right AR | .001 (.062) | .051 (.070) | .000 (.004) | [-.018, .032] |
|  | Between Errors | Left AR | .001 (.086) | -.013 (-.020) | .000 (-.002) | [-.030, .035] |
|  |  | Right AR | .003 (.254)* | .064 (.088) | .000 (.022) | [-.026, .086] |
|  | Strategy | Left AR | -.001 (-.025) | -.009 (-.014) | .000 (.000) | [-.025, .030] |
|  |  | Right AR | .003 (.072)* | .049 (.067) | .000 (.005) | [-.016, .036] |
| **BCS** |  |  | *B*(β) | *B*(β) | *B*(β) | 95%CI |
|  | GEC | Left AR | .001 (.078) | -.475 (-.027) | -.001 (-.002) | [-.028, .019] |
|  |  | Right AR | .001 (.062) | 1.154 (.061) | .001 (.004) | [-.022, .027] |
|  | Between Errors | Left AR | .001 (.086) | -.773 (-.044) | -.001 (-.004) | [-.033, .025] |
|  |  | Right AR | .003 (.254)* | 1.639 (.086) | .005 (.022) | [-.033, .084] |
|  | Strategy | Left AR | -.001 (-.025) | -.652 (-.037) | .001 (.001) | [-.019, .027] |
|  |  | Right AR | .003 (.072) | 1.177 (.062) | .003 (.005) | [-.015, .041] |
| **CRR** |  |  | *B*(β) | *B*(β) | *B*(β) | 95%CI |
|  | GEC | Left AR | .001 (.078) | .191 (.027) | .000 (.002) | [-.018, .032] |
|  |  | Right AR | .001 (.062) | -.376 (-.050) | .000 (-.003) | [-.034, .019] |
|  | Between Errors | Left AR | .001 (.086) | -.053 (.008) | .000 (.001) | [-.027, .035] |
|  |  | Right AR | .003 (.254)* | .-.362 (-.048) | -.001 (-.012) | [-.081, .050] |
|  | Strategy | Left AR | -.001 (-.025) | .072 (.010) | .000 (.000) | [-.020, .028] |
|  |  | Right AR | .003 (.072) | -.417 (-.055) | -.001 (-.004) | [-.039, .019] |

*Note.* Mediation analysis on the association between executive control (Global Executive Composite [GEC] scores of the BRIEF- A, Between Errors and Strategy scores of the Spatial Working Memory task [SWM]) and Coping styles and the mediating role of left and right amygdala reactivity (AR). Gender, age, and the level of education were entered as covariates (*N* = 137). Higher GEC and SWM scores indicate a greater level of dysfunction. See figure 2 for a schematic overview of the associations.

* significant at the .05 level after Holm-Bonferroni adjustment for multiple comparisons. Confidence Intervals remain uncorrected for multiple comparisons.

**Table S3** *Moderation Effect of Group on the Association Between Executive Control and Coping*

|  |  | PAS | |  | CRR | |  | BCS | |
| --- | --- | --- | --- | --- | --- | --- | --- | --- | --- |
|  |  | *B* | *p* |  | *B* | *p* |  | *B* | *p* |
| GEC *Group |  | .001 | 1.000 |  | -.009 | 1.000 |  | -.287* | .009* |
| BE*Group |  | -.001 | 1.000 |  | .001 | 1.000 |  | .005 | 1.000 |
| ST*Group |  | .003 | 1.000 |  | -.007 | 1.000 |  | -.022 | 1.000 |

*Note.* Moderation analysis on the effect of group (patient/control) on the association between executive control (Global Executive Composite [GEC] scores of the BRIEF-A, Between Errors and Strategy scores of the Spatial Working Memory task [SWM]) and coping (Positive Appraisal Style [PAS], Corona-Related Reappraisal [CRR], and the Behavioral Coping Scale [BCS]). Gender, age, and the level of education were entered as covariates (*N* = 137). Higher GEC and SWM scores indicate a greater level of dysfunction.

*significant at the .05 level after Holm-Bonferroni adjustment for multiple comparisons.
